# Predicting progression-free survival in glioblastoma: influence of the perilesional oedema and white-matter disconnectome

**DOI:** 10.64898/2026.02.23.26345834

**Authors:** Manaal Tariq, James. K. Ruffle, Morag Brothwell, Samia Mohinta, Ashviniy Thamilmaran, Honey Panchal, Michael Kosmin, Naomi Fersht, Sebastian Brandner, Parashkev Nachev, Harpreet Hyare

## Abstract

**Background:** Glioblastoma (GBM), Isocitrate dehydrogenase-wildtype (IDH-wt) is characterised by diffuse infiltration, with progression often arising from perilesional tissue and occult white-matter damage. We investigated whether radiomics from the T2/FLAIR-defined oedema and the structural disconnectome improve prediction of progression-free survival (PFS).

**Methods:** We retrospectively analysed 387 adults with newly diagnosed GBM, IDH-wt treated at a single tertiary centre (2005–2020). A deep-learning pipeline segmented enhancing tumour, non-enhancing tumour, and oedema on pre-operative MRI; lesion masks were propagated to normative tractography to derive disconnectome maps. 3-D shape radiomic features extracted for each segmented region underwent appropriate feature selection. Finally, 10 tumour and 9 oedema radiomics were combined with 6 clinical features to train 3 survival models (Random Survival Forest (RSF), XGBoost, Cox proportional hazards (CPH)) that were evaluated on a held-out 20% test set using Harrell’s C-index, Kaplan–Meier risk stratification and time-dependent ROC curves.

**Results:** The best performance was achieved by RSF using all clinical and radiomic features (C-index 0.665 vs 0.595 for clinical features only, p=0.088). Models including oedema radiomics outperformed those using tumour radiomics alone, and disconnectome features, derived from both tumour and oedema regions, were repeatedly selected among the top predictors across algorithms. Combining radiomic and clinical features improved risk stratification and 12-month early-versus-late recurrence classification (AUC 0.704 vs 0.582 for clinical features alone).

**Conclusions:** Integrating perilesional oedema and white-matter disconnectome MR features with clinical and molecular data enhances prediction of PFS in GBM, IDH-wt. These network-aware, multimodal survival models may support personalised risk-adapted treatment strategies pending external validation.

**Key Points:**

- GBM IDH-wt exhibits a high recurrence rate despite aggressive treatment.
- Addition of high-dimensional oedema and disconnectome radiomic features to clinical features showed consistent improvement in the test performance of 3 ML models.
- This can support informed clinical decision-making.

**Importance of the Study:** Prediction of progression free survival (PFS) for a patient with highly recurrent glioblastoma IDH-wt traditionally relies on clinical history, demographics, and molecular markers of the tumour. Recent literature reveals the tumour’s disruptive nature through its invasion of white-matter tracts and identifies its microenvironment, particularly the perilesional oedema, as a harbour of treatment resistant tumour cells. This study is the first to combine high-dimensional radiomic features of the tumour, the oedema, and their disconnectome with clinical and treatment factors to predict PFS. Using 3 model architectures (XGBoost, RSF, and CoxPH), we demonstrate consistent directional improvements in performance, on addition of radiomic features to clinical baseline models. Furthermore, oedema and disconnectome radiomics are identified as top predictor features across algorithms. This proof-of-concept study provides a reproducible multimodal pipeline, reaffirms the usability of MR radiomics, and identifies features of the oedema and the structural connectome as promising biomarkers, demanding large-scale external validation.

## Introduction

Glioblastoma (GBM) is the most common and lethal primary malignant brain tumour in adults, with a median survival of approximately 15 months despite maximal safe resection, radiotherapy (RT), and chemotherapy. ^1^ Its infiltrative nature leads to high rates of local recurrence and limited progression-free survival (PFS). The 2021 WHO Classification now defines “glioblastoma, IDH-wt” (CNS WHO Grade 4) as a distinct entity, characterised histologically by necrosis and/or microvascular proliferation or, in certain cases, molecular markers such as telomerase reverse transcriptase (TERT) promoter mutation, epidermal growth factor receptor (EGFR) amplification or combined +7/–10 copy number change. ^2^

Although many efforts have focused on the enhancing tumour core for prognosis and treatment planning, mounting evidence highlights the importance of the perilesional region — in particular, the T2/FLAIR-hyperintense perilesional oedema zone — as a substrate for early recurrence. For instance, radiomic studies indicate that heterogeneity within the perilesional oedema is associated with shorter PFS and is predictive of the site of recurrence in GBM.^3^ This suggests that the margin of the lesion — often bypassing gross resection — may harbour radio- and chemo-resistant tumour-infiltrative cells or a permissive microenvironment.

Moreover, the concept of the disconnectome, ^8^ which represents the structural white-matter network disruption caused by tumour infiltration and oedema, is increasingly shown to carry prognostic information. Quantitative assessments of structural connectome disruption in GBM patients reveal that lesions cause far-reaching white-matter changes beyond the visible mass, and that the degree of structural connectivity disruption is associated with worse survival outcomes.^4^ Using high-dimensional radiomic features to capture changes in the tumour morphology, perilesional extent, and white-matter disruption forms a multimodal, network-aware prognostic approach.^5^

Building on these insights, the present study investigates whether 3-D shape-based radiomic features of the original lesion mask and the disconnectome, derived from the full tumour and perilesional oedema, in combination with clinically relevant variables and molecular markers (MGMT methylation, TERT mutation) can improve machine-learning (ML) based prediction of PFS in IDH-wt GBM patients. We hypothesise that integrating these quantitative imaging biomarkers with clinical/molecular data will provide a more generalisable and interpretable prediction of time to progression, thus supporting personalised decision-making and treatment stratification.

## Materials and methods

### Study Population

969 consecutive patients with a histological diagnosis of IDH-wt GBM treated at National Hospital for Neurology and Neurosurgery (NHNN) between 2005 and 2020 were identified. ^43^ 105 patients were excluded due to mismatched pathology, 3 due to duplication, and a further 474 patients were excluded due to lack of follow-up data. The final cohort consisted of 387 patients, all of whom received RT. Of these, 379 had documented radiological recurrence and 8 had not recurred by the end of the follow-up period. This formed a cohort with a 97.9% event rate and a 2.1% censoring rate, deemed appropriate for the study’s focus. Figure 1 demonstrates the step-by-step flow of data selection leading to the formation of the study cohort.

**Figure 1:**
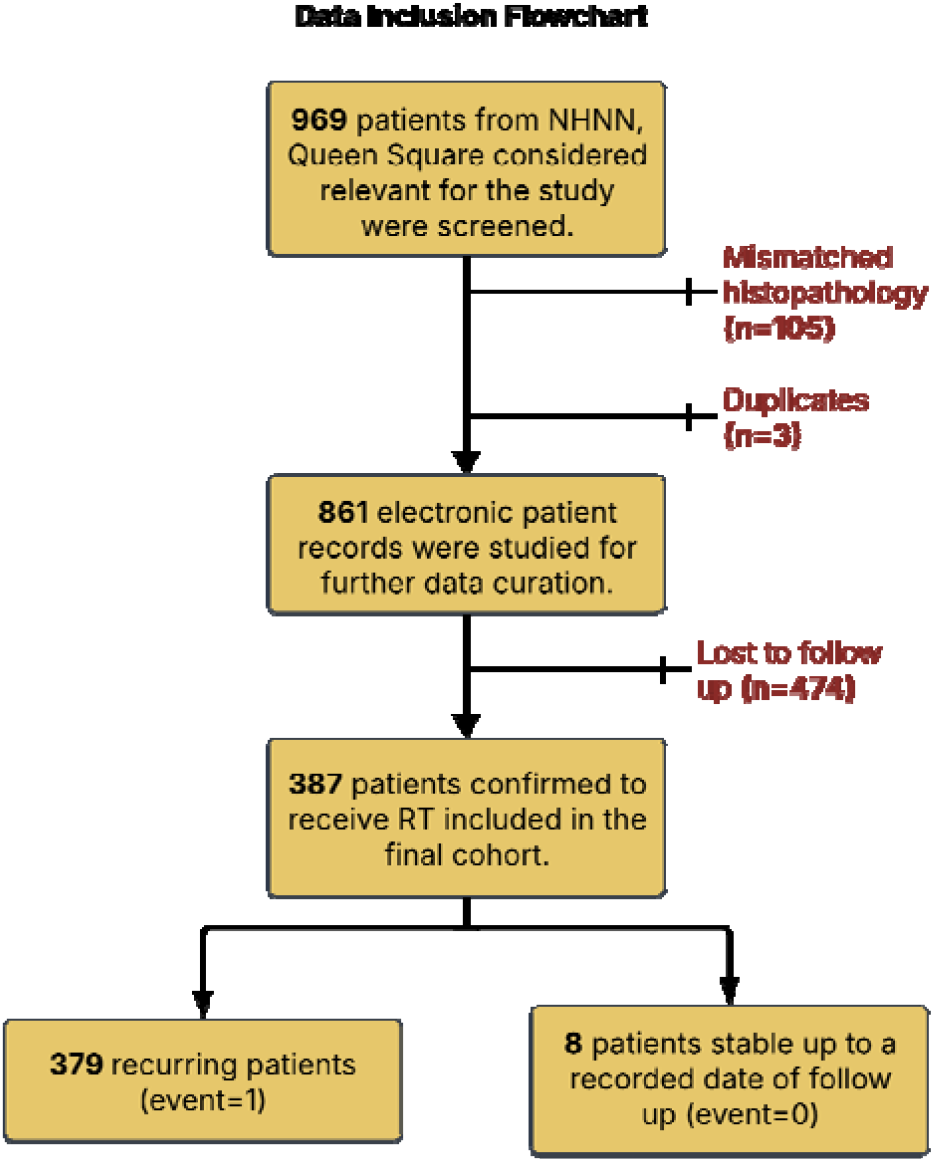
Flow of patient inclusion/exclusion from the final dataset used in the study (n=387). All patients had undergone RT and either had a defined PFS (n=379) or a duration of no recurrence (n=8) up till the end of their follow up period.

Data was carefully extracted to account for patient history (age, sex, birth, and death dates), diagnosis (histopathology, key molecular markers, diagnostic and recurrent MRI), and treatment (date and type of surgical intervention, RT dose and duration). These were used to calculate time from operation to progression, time from the end of RT to progression, and overall survival in days. For this study, PFS is defined as the time from operation (start of treatment) to first radiologically confirmed recurrence, based on the Response Assessment in Neuro-Oncology (RANO) criteria and evidence of an increase in mass effect or tumour size on MRIs (Figure 2). ^6^ MRI’s used included MRI Head/Head + Contrast and MRI Stealth/neuronavigation. Data were acquired on multiple scanners across the NHS Trust.

**Figure 2:**
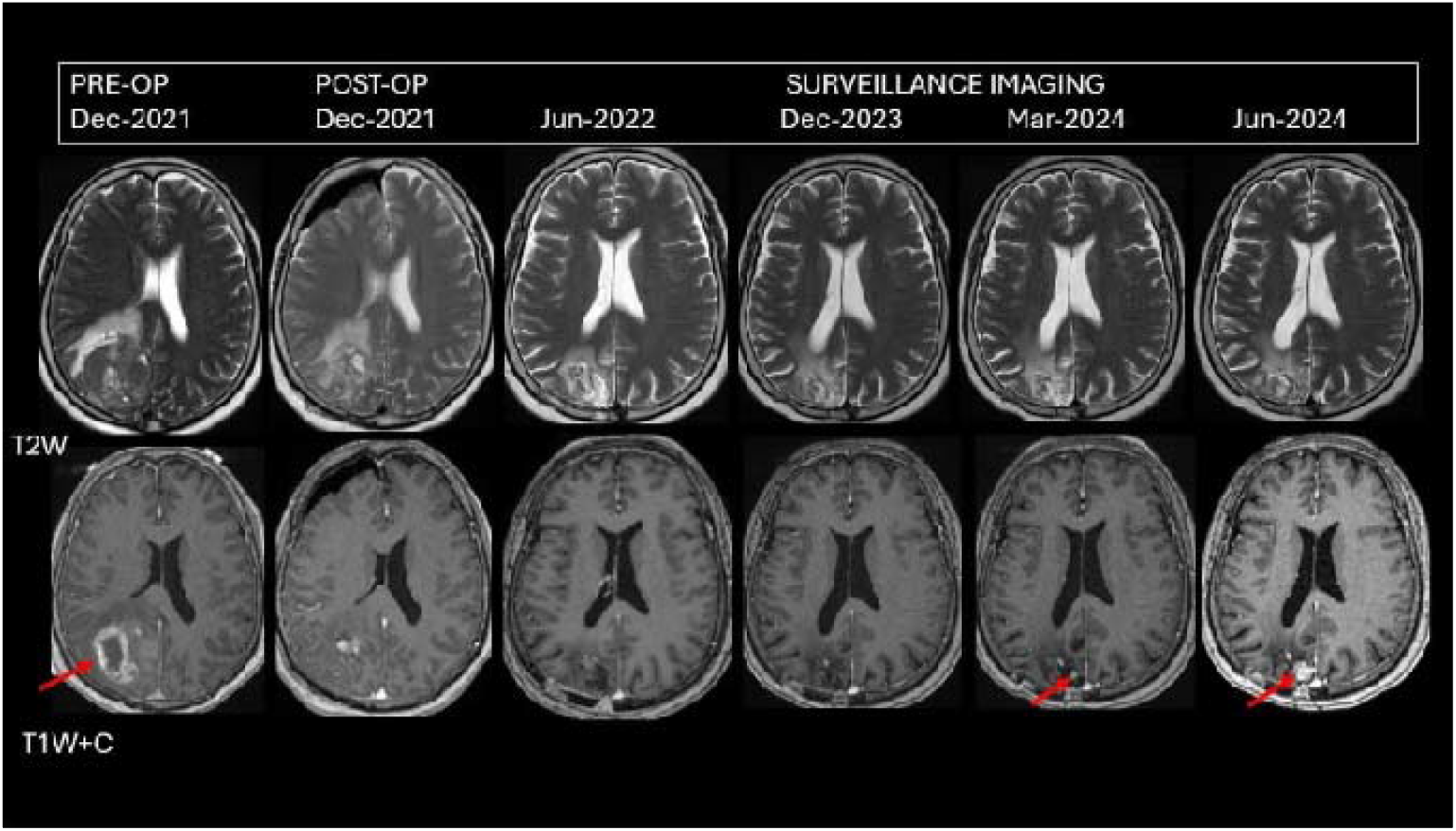
Longitudinal imaging of a patient in their early 50s with GBM IDH-wt. T1-weighted contrast-enhanced (T1CE) image sequence shows the pre-operative tumour bulk in December 2021, followed by macroscopic excision. Subsequent imaging reveals mass recurrence (as highlighted by the red arrow) in March 2024. The corresponding T2-weighted sequence shows persistent hyperintensity around the mass, consistent with the perilesional oedema, which remains stable across timepoints.

Operative procedures included either biopsy or debulking (where excised tissue was used for biopsy). Follow-up surgical procedures, such as shunt revisions or repeated debulking, were not recorded, as the study aimed to predict the first recurrence. Radiotherapy regimens varied, with most patients receiving Intensity-Modulated Radiation Therapy (IMRT), often with concurrent and adjuvant temozolomide. A few patients had either missing radiotherapy doses despite completing the course or dates of death. Intermittent records can be attributed to hospital/country/hospice transfers, rapid progression, or treatment refusal. Data missingness was accounted for in our data pre-processing pipeline (see below).

### Segmentation Pipeline

A fully automated, deep learning based in-house segmentation pipeline was used for tumour segmentation. ^7^ GBMs were segmented into three regions (oedema, non-enhancing tumour, enhancing tumour), with the enhancing and non-enhancing tumour together referred to as the full tumour and all sub-regions coming together to form the full lesion. The segmented lesions were then used to create corresponding disconnectome maps, as shown in Figure 3.^8^

**Figure 3:**
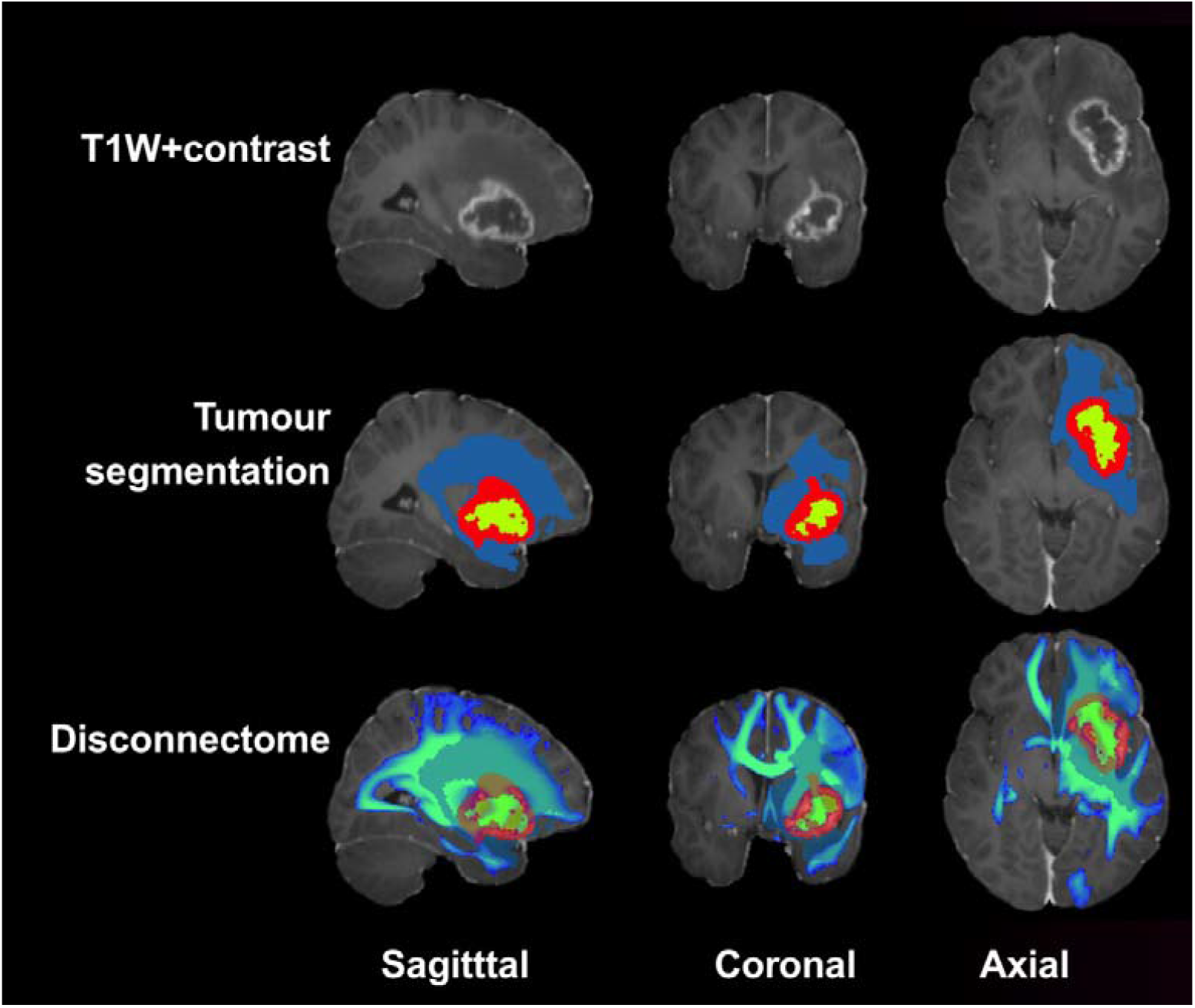
Top panel shows the T1-weighted post contrast series of a patient in their late 30s with a glioblastoma. Middle panel demonstrates tumour segmentation output: green non-enhancing, red enhancing, and blue oedema. Bottom panel further maps the estimated disconnectome using this lesion segmentation. ^7^

These segmented GBMs and disconnectome maps were subjected to Pyradiomics-based derivation of 17 three-dimensional shape-based radiomic features for each of the five sub-regions within the tumour, oedema, and their disconnectome. ^9^ A total of 170 radiomic features were obtained (17 lesional and 17 disconnectome for each of the 5 compartments; see Supplementary Table 1).

### Data Preprocessing

The dataset was pre-processed before feature selection and model training to ensure compatibility with ML algorithms. Preprocessing and hyperparameter optimisation were performed exclusively using the training data to prevent data leakage. We used a nested validation strategy: an outer stratified 80/20 split creating training (n = 309) and test (n = 78) sets. Within the training set, 5-fold stratified cross-validation was used for hyperparameter optimisation. Numerical features included all radiomics (full tumour and oedema) and age, categorical data type was used for RT dose (30, 40, 40.05, 54.9, 59.4, 60 Grey (Gy)), sex (M/F) and surgery type (debulking/biopsy). Molecular genetics data on TERT gene amplification and Methylated-DNA-Protein-Cysteine Methyltransferase (MGMT) gene methylation status was binarised (1/0) and treated as categorical data. Numerical data was standardised using the sklearn StandardScaler, yielding z-score normalisation to ensure comparable scales, while categorical information was encoded using LabelEncoder to one-hot encode numerical labels.

Age, RT type, PFS, OS, time from RT to progression, and molecular markers were features missing at random for a few patients (less than 9.8% for all). Missing values in continuous data were handled using median imputation, and in categorical data (RT type, molecular markers) using mode-based imputation. No patient had missing radiomics, surgery, or sex. Trained imputers, encoders, and scalers were then applied to the test set.

### Feature Selection

Six clinical features were used for all models: age, sex, surgery, RT type, TERT mutation, and MGMT methylation status. These were selected *a priori* as directly clinically relevant and were not subject to a feature selection function.

A multi-step feature selection pipeline, summarised in Figure 4, was implemented to select radiomics using only the training set (n = 309) before hyperparameter tuning. The features were only selected once and used across all cross-validation folds before running the first survival model (XGBoost) for computational efficiency and consistency. While variance filtering was used to identify features with low variance (below the 0.01 threshold) across all training samples, hierarchical clustering-based correlation analysis identified highly correlated (Pearson’s |r| > 0.85) pairs and retained the feature with the highest variance within each pair to reduce multicollinearity.

**Figure 4:**
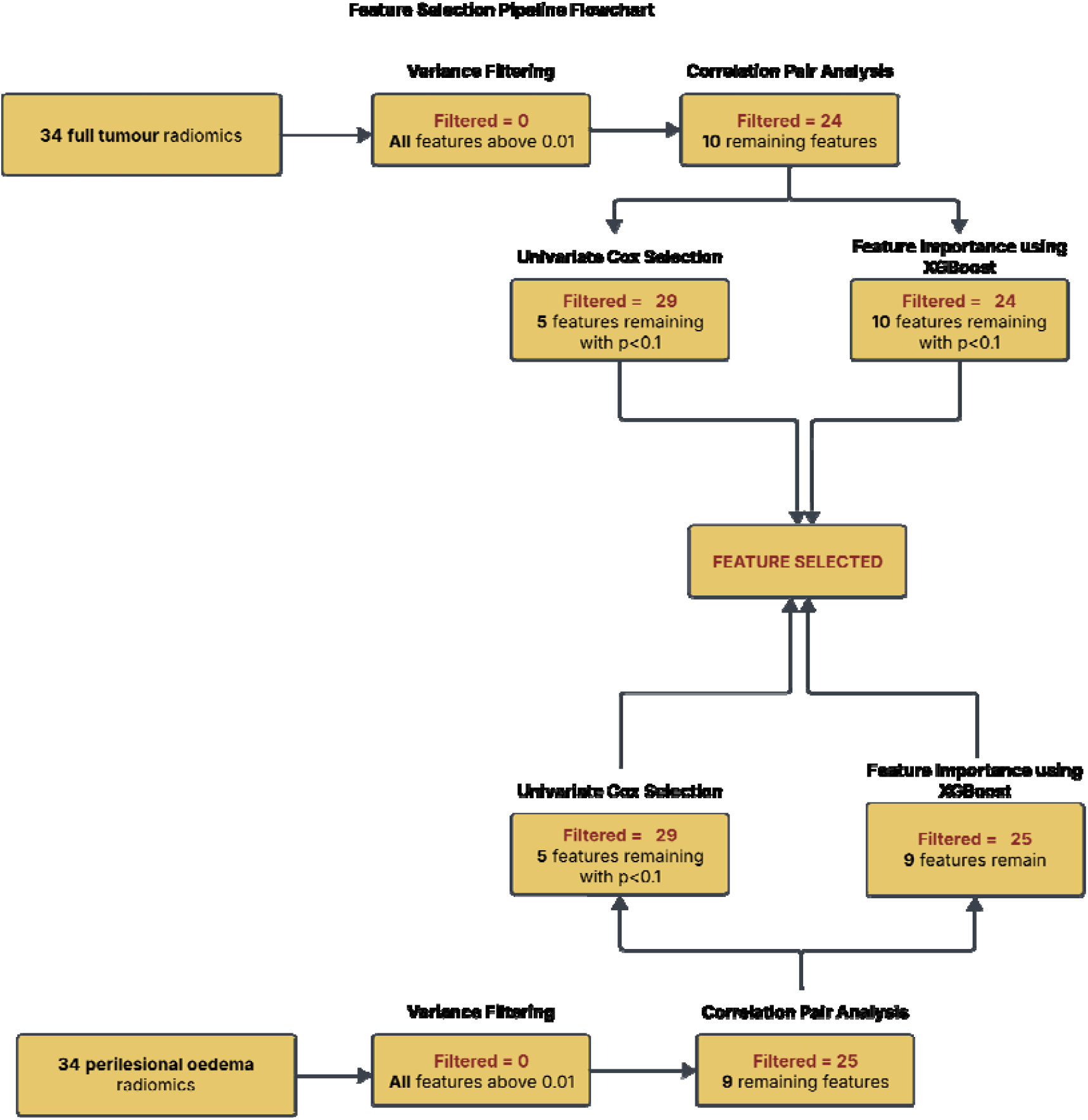
Schematic overview of the sequential multi-step feature selection pipeline applied independently to both tumour and perilesional oedema radiomics. No feature was eliminated post variance filtering, correlation pair analysis removed all but 10 features for full tumour and 9 features for oedema. Independent feature selection methods-univariate Cox proportional hazards (p < 0.1) and XGBoost-based feature importance (top-K by gain) were applied to the features remaining after correlation filtering. Radiomic features identified by either of these two methods were retained using a union-based (OR) selection rule, whereby each method contributed a binary vote and features with at least one vote were selected. No weighting or ranking were applied based on vote count. As a result, the pipeline finally selected 10 tumour radiomics and 9 oedema radiomics.

The subset of features that passed both variance and correlation-based filtering was tested independently in parallel using two methods: univariate Cox regression filtering and XGBoost (gradient boosted tree-based survival model) feature importance selection. The univariate Cox identified statistically significant radiomics with a Wald test p-value <0.1, and the XGBoost importance-based selection ranked features by gain-based importance, retaining up to 15 or all remaining features, whichever was fewer. Radiomic features identified by either method were retained for subsequent modelling, implementing a hybrid filter-wrapper selection strategy. Feature selection was performed exclusively on training data to avoid information leakage.

The pipeline selected 10 tumour features and 9 oedema features, giving us the following feature sets for our models: clinical only (n=6), clinical + full tumour (n=16), clinical + oedema (n=15), and all features together (n=25).

### Model Development and Optimisation

The train/validation set (n=309) was used for feature selection, model training, and hyperparameter tuning whereas the test set (n=78) was only used for final model evaluation to reflect the model’s ability to generalise to unseen data.

We fitted three survival models to predict progression free survival (PFS) (in days) on all four feature sets: XGBoost with Cox objective, Random Survival Forest, and Cox Proportional Hazards. The models were implemented using Python libraries: lifelines (version = 0.28.0), xgboost (version = 1.60), and scikit-survival (version 0.24.1). ^10, 11, 12^

#### 1. XGBoost with a Cox loss function

XGBoost is a fast gradient boosting technique that allows processing of right-censored survival data and uses better approximation of a partial likelihood function as learning objective. ^13^ It uses sequentially boosted trees to model the hazard function of a patient which is the linear function of a population baseline hazard.^14^ The main objective of XGBoost was to model PFS analysis using the survival:cox objective.

#### 2. Random Survival Forest (RSF)

RSF is a non-parametric method that works on analysing right-censored survival data using an ensemble of tree-based learners. ^15^ It is an extension of the Random Forest method and uses a bootstrap aggregation method (sampling method that allows repeated use of the same sample). It makes no assumption about proportional hazards and uses log-rank splitting to separate survival curves. ^16^

#### 3. Cox Proportional Hazards (CoxPH)

The Cox model is a semi-parametric regression model that focuses on maximising the Cox partial likelihood, a classical survival analysis approach. It is a prediction of the hazard rate ratio which is indicative of instantaneous risk between two groups rather than cumulative risk. ^17^

For all 3 models, automatic Optuna hyperparameter technique was implemented to enable sampling and testing of optimal hyperparameter values within a defined range for each model.^18^ It employs a Bayesian optimisation-based algorithm called Tree-structure Parzen Estimator (TPE) which iteratively modeled the behavior of the objective function across 100 trials with 5-fold stratified cross-validation on the train/validation set (n=309, 80%) and each fold using approximately 247 patients for training and 62 for validation. ^19^

Optimal hyperparameters were selected based on the highest mean cross-validation C-Index. Adaptive regularisation was implemented for each of the four feature sets to ensure hyperparameter constraints were stricter for models with more features to avoid overfitting and to improve model generalisation (see supplementary table 2). Hyperparameter combinations that showed excessive overfitting were penalised (train-validation C-index difference >0.15 for XGBoost and >0.10 for RSF) to ensure generalisability.

### Model Evaluation and Validation

Harrel’s Concordance index (C-Index), with 95% confidence intervals computed across 1000 bootstrap resamples (seed = 42 for reproducibility), was used to evaluate predictive performance of all three survival models as they perform time to event analysis. C-index quantifies the correlation on a scale of 0-1, where higher values reflect better concordance between actual observed time to event and its predicted value, distinguishing between higher and lower risk (earlier vs later recurrence). ^20^ Statistical significance was evaluated at *α* = 0.05 through pairwise model comparisons using paired bootstrap hypothesis testing.

All models were used to create corresponding feature importance plots: 1) Shapley Additive exPlantations values are calculated using the XGBoost model predictions and corresponding values for each sample are plotted to understand directionality and impact on model outcome. 2) Random Survival Forest is a model-agnostic method that uses a permutation-based method to evaluate the most important features. Random permutation of feature values increases model prediction error if the feature is more important, hence allowing feature ranking. 3) For the semi-parametric CoxPH model, we create a plot that visualises hazard ratios *exp*(*β*) which are estimates of the ratio of hazard rates or instantaneous risk between two groups calculated by the model coefficients (*β*). ^17,21^

Kaplan Meier curves were generated using the best performing model’s predictive risk scores that allowed classifying test set patients into high, medium, and low risk groups for disease progression/recurrence. Log-rank test was used to assess statistical significance and monotonic ordering (low > medium > high time to recurrence) was evaluated for appropriate risk group calibration.

Furthermore, time-dependent Receiver Operating Characteristic (ROC) Curve analysis at 6, 12, 18, and 24 months was performed using model predicted risk scores to evaluate the best performing models’ ability to predict early versus later recurrence in patients who recurred within the test set (n=76). Censored patients within the test set (n=2) were excluded as they did not experience recurrence. Patients were described to have early recurrence if they recurred within the selected time frame (6/12/18/24 months) and late recurrence if they recurred post that time frame or remained recurrence free. The evaluation parameter is Area Under Curve (AUC) which discriminates between the binary outcomes at various threshold settings. ^12^

## Results

### Patient Cohort

The study cohort comprised 387 patients confirmed as GBM, IDH-wt (no mutation). Among these, 97.9% had confirmed disease recurrence during follow-up, and 93.5% were deceased at the time of data lock. In terms of initial treatment, 71.3% patients underwent debulking surgery, while the remaining underwent biopsy procedures. All patients received radiotherapy; however, 93.3% had details of the exact dose available in their records. Table 1 summarises the distribution of the clinical dataset.

**Table 1:** Clinical Summary of Curated Patient Dataset. IQR is short for inter-quartile range.

| STUDY COHORT |  |  |
| --- | --- | --- |
| SAMPLE | n = 387 | Data availability % |
| AGE | 59 (IQR 50.0–67.0) | 99.2% available |
| SEX | 245 Male, 142 Female | 100% available |
| SURGERY TYPE | 276 debulking, 111 biopsies | 100% available |
| RADIOTHERAPY TYPE (RT TYPE) | 30 Gy, 6# (n= 53)<br>40-40.05 Gy, 15# (n = 14)<br>54-54.9 Gy, 27-30# (n = 18)<br>59.4 (33#) - 60 Gy (30#) (n = 264)<br>Miscellaneous Dose (n = 12)<br>Dose Missing (n = 16) | 93.3 % available |
| TIME FROM OPERATION TO PROGRESSION (PFS IN DAYS) | n = 379<br>274 (IQR 153.0–442.5) | 97.9% available |
| TIME FROM END OF RT TO PROGRESSION | n = 370<br>184.5 (IQR 75.5–345.5) | 95.6% available |
| OVERALL SURVIVAL (DAYS) | n = 353<br>501, IQR (344.0–758.0) | 91.2% available |
| MOLECULAR MARKER (MGMT METHYLATED) | 93 MGMT methylation, 293 no MGMT methylation | 99.7% available |
| MOLECULAR MARKER (TERT MUTATION) | 103 TERT mutated, 283 not TERT mutated | 99.7% available |

### Model Performance and Feature Importance

We assessed the performance of three ML survival models by analysing their predictive accuracy through C-Index values on the held-out test set (n=78, events=76).

The models were trained on four feature sets: clinical only, clinical + tumour, clinical + oedema, and all features together as summarised in figure 5 (A). All three survival model types demonstrated a consistent increase in performance on adding radiomics to the clinical-only model with the clinical + oedema (n=15) feature set performing better than the clinical + full tumour (n=16) feature set for both RSF and CoxPH.

**Figure 5:**
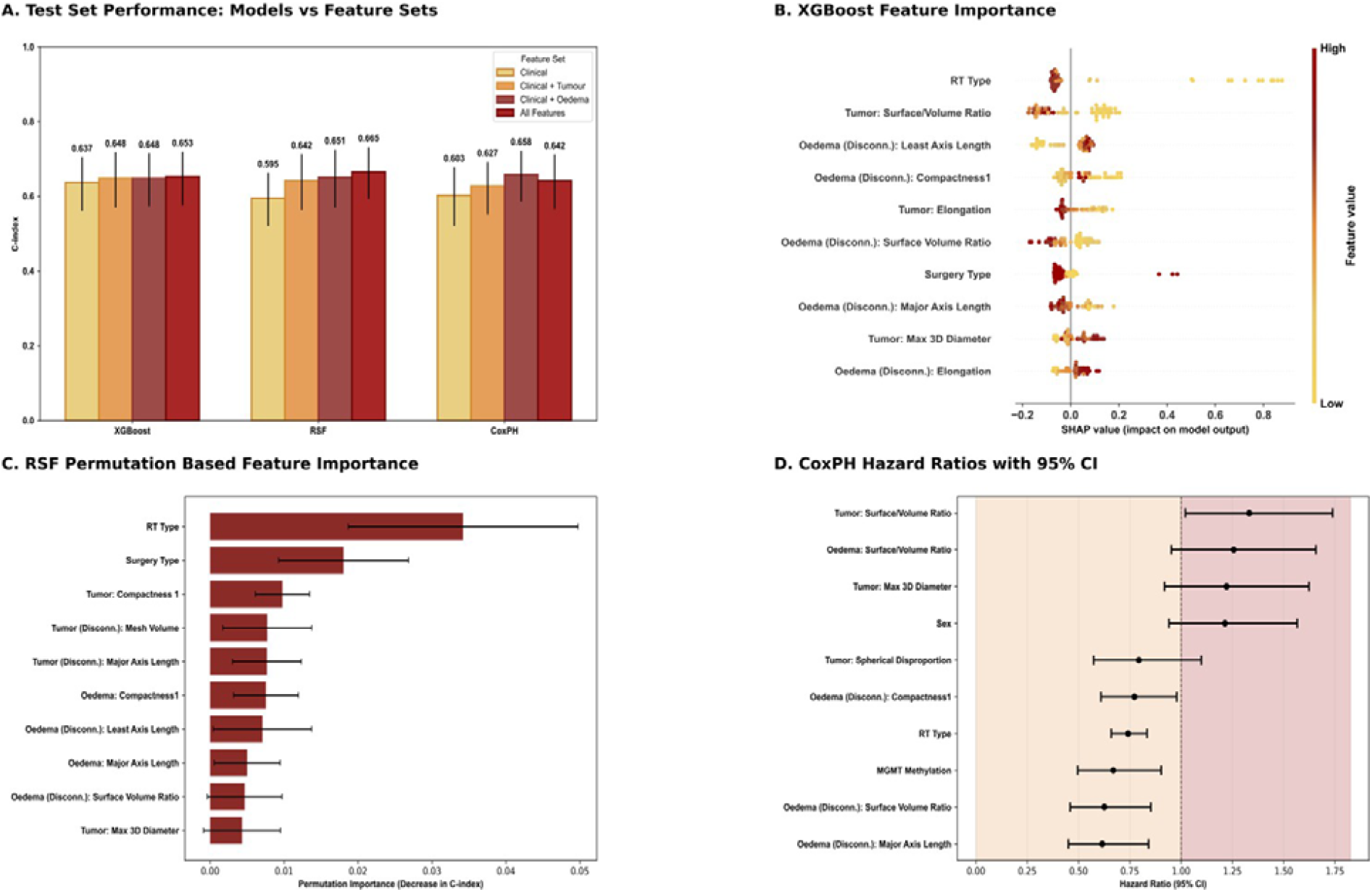
Model Performance and Feature Importance A) C-Index across XGBoost, RSF, and CoxPH models evaluated on held-out test set (n=78) for all four feature sets (clinical, clinical + tumour, clinical + oedema, all features). B) XGBoost feature importance using SHAP values calculated based on predicted risk scores ranked from most to least influential. Each dot represents an individual patient in the test set (n= 78), with their colour indicating feature value (red= high, yellow = low). The x-axis shows SHAP value magnitude where positioning on the right implies a greater risk of recurrence and to the left suggests lower risk. C) Feature Importance for RSF ranks from most to least impactful based on highest to lowest decrease in C-index following feature permutation. D) CoxPH feature importance, categorised by direction and magnitude of effect. Values < 1 (orange) indicate reduced risk, while values >1 (red) indicate increased risk.

The best results were achieved on the RSF model trained on all clinical and radiomic features, which attained a C-Index of 0.665 (95% CI: 0.593-0.732), compared to the clinical only model with a C-Index of 0.595 (95% CI: 0.521-0.663). A similar trend was observed on the fitted XGBoost model where the model trained on all features (C-index 0.653, 95% CI: 0.575-0.719) outperformed clinical-only (C-Index 0.637, 95% CI: 0.561-0.705) and CoxPH (clinical-only C-index: 0.603 and all-features with a C-index of 0.642). Although individual pairwise comparisons did not achieve statistical significance (all features vs clinical-only for RSF (Δ = 0.0709, *p* = 0.088), for XGBoost (Δ = 0.0156, *p* = 0.73), and for CoxPH (Δ = 0.0413, *p* = 0.306)), the directional improvement in performance on feature addition across all models is indicative of the prognostic value of multimodal radiomic signatures.

Feature importance analysis corresponding to each model highlighted the top 10 of 25 features for each of the 3 algorithms trained on all features. Figure 5 (B) summarises the results of the XGBoost SHAP analysis which identified 2 clinical, 3 tumour, and 5 oedema features with mean SHAP values of 0.160, 0.120, and 0.079 respectively for the top 3 features. Mean SHAP value is representative of the feature’s impact on average shift in the log-risk score.

RSF permutation-based analysis highlighted 2 clinical, 4 tumour, and 4 oedema features in figure 5 (C), demonstrating how permutations of the listed feature’s values contributed to the change in C-index. Feature permutations caused a decrease of 0.0342 ± 0.0155 in C-index for RT type, 0.0180 ± 0.0088 for surgery type, and 0.0098 ± 0.0037 for compactness of tumour. The lowest reduction in C-Index (0.0043 ± 0.0052) among the 10 was caused by maximum 3D diameter of the tumour.

Figure 5 (D) arranges the top 10 features (3 clinical, 3 tumour, and 4 oedema features) for CoxPH model by the magnitude and direction (protective vs risk-enhancing) of their hazard ratios. It does not follow the same ranking system as the other two methods. The most statistically significant feature for the model is RT type with a hazard ratio of 0.741 (95% CI: 0.659-0.834, p=0.00) indicating a 25.9% lower risk of recurrence compared to patients receiving lower doses. It is followed by the following three features: MGMT methylation status (HR=0.669, 95 % CI: 0.496-0.903, p=0.0086), and major axis length (HR=0.615, 95% CI:0.450-0.841, p=0.0024) and surface volume ratio (HR=0.626, 95% CI: 0.459-0.852, p=0.0029) of the oedema disconnectome.

RT type has the highest impact on both XGBoost and RSF model performance and the most significant impact on the CoxPH model performance, demonstrating generalisability across model types.

### Survival plots

We compared each of the four feature sets used to model survival using the best performing algorithm, *RSF*, by creating Kaplan Meier survival risk curves (Fig 6).

**Figure 6:**
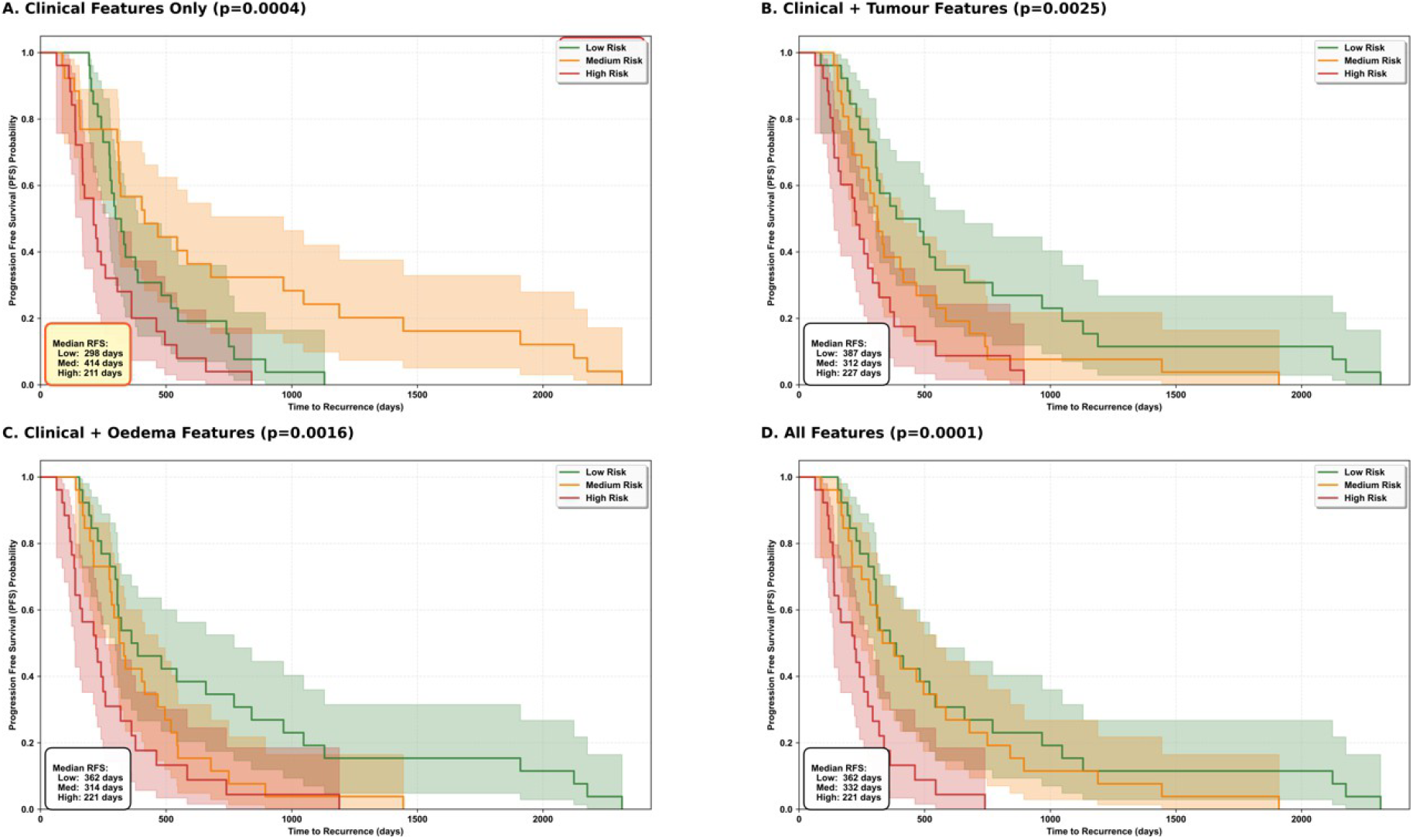
Kaplan Meier curves made using RSF models trained on the test set (n=78) for each of four feature sets (clinical, clinical + tumour, clinical + oedema, all features) demonstrating statistically significant risk based stratification for all feature sets except clinical.

The clinical-only model showed non-monotonic risk group ordering, with median-risk patients showing longer median survival of 414 days compared to low-risk patients with median survival of 298 days. Despite a significant log-rank test p value (0.0004), this model performs the worst amongst the four feature sets due to poor model calibration as also indicated by its C-index value (0.595). Models with radiomics showed monotonic risk-group ordering and performed better than the clinical only model.

The best separation is produced when patients were classified by low, medium, and high risk using the RSF model trained on all clinical and radiomic features (p=0.0001) followed by clinical + oedema features (p=0.0016), and finally clinical + tumour features (p=0.0025).

For the best performing feature set (all features), low-risk patient group has a median recurrence time of 362 days, medium risk group has a median recurrence time of 332 days, and high-risk group has a mean recurrence time of 221 days.

### Early versus later recurrence classification

The best performing model (RSF) was used to create ROC curves at thresholds of clinically relevant timepoints of 6, 12, 18, and 24 months for recurring patients within the test set (n=76) (Fig 7).

**Figure 7:**
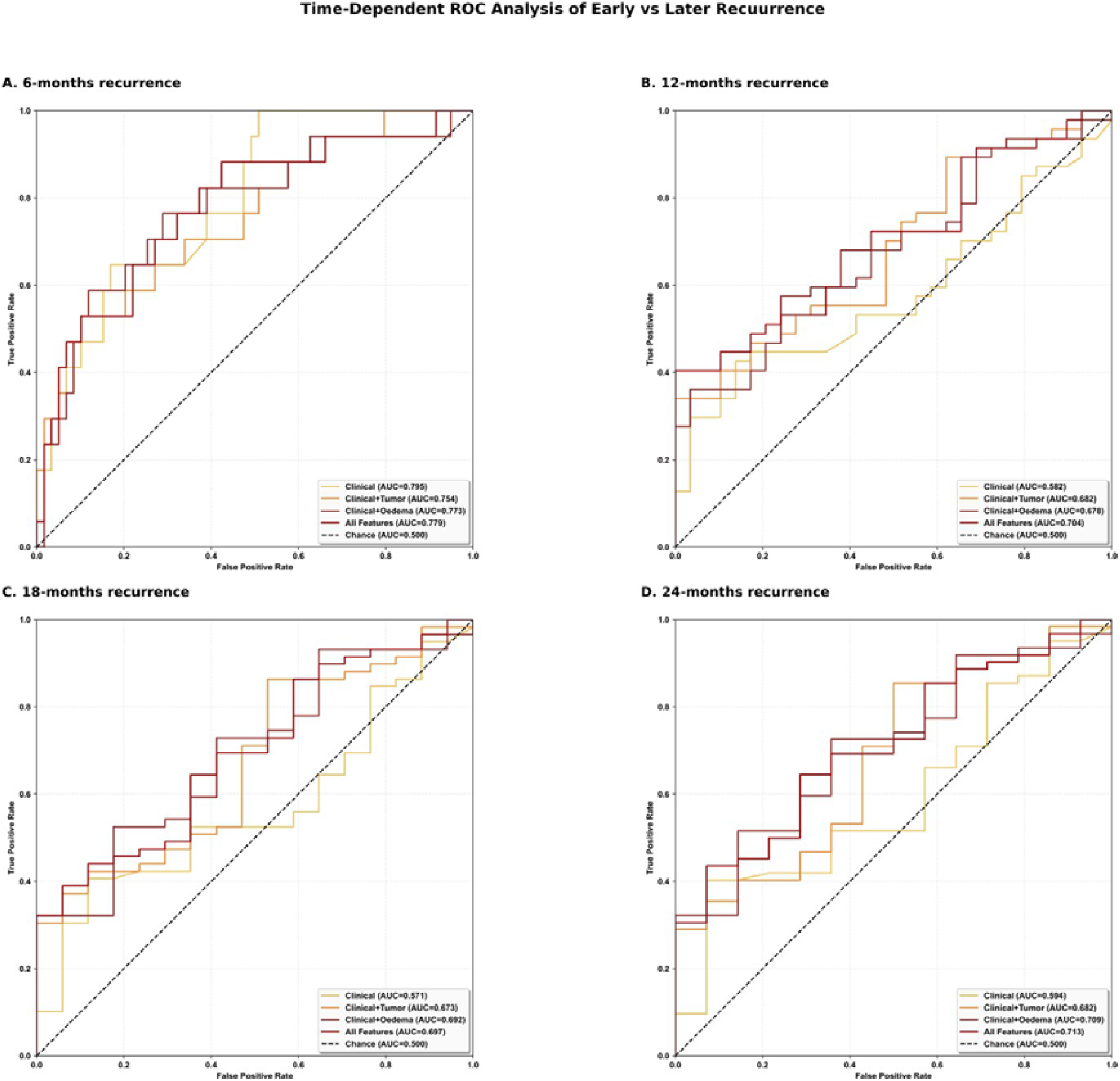
RSF based receiver operative characteristics (ROC) curves for predictive recurrence outcomes at: (A) 6-months, (B) 12-months, (C) 18-months, (D) 24-months recurrence. Splits of samples into early: late recurrence were 17:59 for (A), 47:29 for (B), 59:17 for (C), and 62:14 for (D).

At 6 months, highest discriminative ability was achieved by clinical features alone (AUC:0.795) whereas All Features outperformed clinical features alone at 12 (AUC: 704), 18 (AUC:697), and 24 months (AUC: 713). ROC analysis was limited at 6 months by the small number of early recurrence events (n=17) compared to 59 post 6-month recurrences, which resulted in reduced statistical power for between-feature set comparisons.

The RSF model was most effective in classifying risk between patients in the test set at 12 months which is the optimal recurrence time point for this patient cohort with a ratio of 47:29 early to late recurrence events.

## Discussion

We have shown that adding quantitative imaging biomarkers from the perilesional oedema and white-matter disconnectome meaningfully improves PFS prediction beyond standard clinical and molecular factors in adult IDH-wt GBM. MRI features derived from the oedema and its disconnectome not only outperformed tumour-only features but also dominated the top-ranked predictors across XGBoost, RSF and CoxPH importance analyses, highlighting the prognostic relevance of the peri-tumoural microenvironment and network disruption. The all-features RSF model enabled well-calibrated risk stratification with clearly separated Kaplan–Meier curves and improved discrimination of early versus later recurrence at clinically relevant timepoints, particularly at 12 months (AUC 0.704 vs 0.582 for clinical-only). Collectively, these findings support perilesional oedema and disconnectome radiomics as key components of multimodal predictive recurrence models for GBM.

Recurrent or progressive glioblastoma occurs in virtually all patients diagnosed with this tumour type and is characterised by the presence of infiltrative tumour cells and even an interaction between the infiltrative and apparently normal cells in the perilesional oedema.^22^ Given its heterogeneity and the interplay of clinical, demographic, and tumour associated features, predicting prognosis becomes a complex task. To address this, our study used three survival model approaches combining clinical, molecular, lesional and disconnectome radiomic features of the oedema and tumour to predict first progression. Progressive addition of radiomics to the baseline clinical only model showed consistent improvement in performance for ensemble tree-based methods (XGBoost, RSF). The ‘All Features’ feature set showed superior prognostic accuracy with a C-Index of 0.653 for XGBoost and 0.665 for RSF The integration of orthogonally derived unimodular data in the form of diverse spatial radiomics with clinical/molecular data is essential for capturing the comprehensive multimodal signature necessary for holistic prognosis prediction. ^23^

The three models demonstrated relative improvements in C-index by 2.5% (XGBoost), 11.8% (RSF), and 6.5% (CoxPH) from the baseline clinical model to the model integrating all features together. However, while the ‘All Features’ feature set performed best for both ensemble methods, it was the ‘Clinical + Oedema’ feature set that outperformed any other for CoxPH. High dimensional features such as radiomics can have counter-intuitive geometric properties that make their behaviour hard to interpret. ^24^ Radiomics of spatially adjacent tumour and oedema regions can exhibit correlation causing signal dilution and reduced discriminative performance. ^25^

Furthermore, this model-dependent feature preference reflects inherently distinct model architectures. While CoxPH is a log-linear model which assumes the hazard ratio between two groups to be constant over time, both ensemble methods (XGBoost and RSF) do not assume this linearity and are thus more suited to capture non-linear relationships such as those between radiomics/clinical variables and recurrence time.^26^ CoxPH prioritises building a more clinically interpretable model whereas XGBoost and RSF prioritise optimising model performance and can better handle feature redundancy through recursive partitioning and feature subsampling. ^15,27^

Our study cohort exhibits a 97.9% event rate that provides sufficient statistical power and is suitable to represent the aggressive nature of the tumour, yet still accounting for patients who did not recur up to a certain time point.^27^ Prognostic studies around PFS and OS in GBM have been focused on binary classification of recurrence/death risk.^29,30,31^ However, survival models simultaneously do two things: decide if a defined event has occurred and estimate when it occurred using censored and uncensored observations.^32^ Given the high event rate, our models are primarily characterising recurrence through calculating a continuous time to recurrence rather than simply classifying whether the tumour has recurred. ^33^

Apart from predicting recurrence, we investigated the top 10 features contributing most to each model trained on ‘All Features’. Repetition of features across the three algorithms highlights generalisability of the feature set and emphasises a robust model. Oedema radiomics formed 50% of the top 10 ranking features for XGBoost and 40% for both RSF and CoxPH. This competitive performance shows the perilesional oedema to be a prognostically important region, possibly due to local tissue disruption and tumour-infiltrative patterns. This is further echoed by the response assessment framework (RANO criteria) which was primarily revised to incorporate and evaluate diffused and infiltrative tumour growth using T2-w and FLAIR MRI sequences.^6^ Furthermore, it reiterates the role of radiotherapy resistant glioma cells, found in the perilesional oedema, in disease recurrence^34,35,36,37^ and its link to reduced OS. ^37^

The prominent role of the white matter disconnectome is underscored by the consistent appearance of its radiomics in the top 10 features across model types (5/10 in XGBoost, 4/10 in RSF, and 3/10 in CoxPH). It supports our network-aware modeling approach and shows PFS to be a function of both local tumour properties and the spatial extent of white-matter tract involvement. Disconnectome radiomics are highly suited to studies focused on IDH-wt tumours which are characterised by a high proliferation rate, destructive capacity, and a disruptive glioma-tract relationship.^39^ Connectome disruptions are associated with worse survival, cognitive function, and their quantitative integration provides a robust prognostic marker of disease progression.^4^

It is important to note that our study concentrates on assessing first recurrence post diagnosis and radiotherapy despite several patients having recurred more than once. First progression for patients with GBM is a critical clinical decision point as per the standardised RANO criteria as it most accurately captures the impact of the initial standard-of-care therapy.^40^ Furthermore, subsequent progression is considered to have limited relation to markers found in the primary resected tumours, the subjects of radiomic derivation in this study.^41^

Given that PFS represents the time to radiological detection of recurrence, it may be influenced by the lesion’s location within functionally eloquent regions. Lesions affecting functionally critical networks would likely have a more evident appearance of symptoms leading to earlier diagnosis in comparison to lesions in silent regions that remain asymptomatic for longer and are usually identified through routine surveillance imaging. Disconnectome features that quantify the spatial extent of damage across the brain network can, to some extent, reflect this detection bias however, standardised clinical and imaging protocols can curb its effects in practice.

In contrast to the RSF baseline clinical model, the RSF ‘All Features’ model performed significant monotonic risk stratification of patients as evident from the rapidly declining high-risk KM curve compared to the low-risk curve (Fig 6). Similarly, a significant incremental gain was observed between the AUC on progressive addition of radiomic features across clinically relevant time points. When using all features for prediction, the trained model is expected to produce correct risk stratification 70.4% (AUC=0.704) of the times at the 12-month time point compared to 58.2% accuracy when clinical features are used alone (Fig 7). Effective risk grouping using this model holds clinical relevance as it is a more decisive method of prognostic stratification— supporting informed consent and enabling earlier intervention. For those at high risk of rapid progression, it could support the patient’s decision to dose-escalated radiotherapy or experimental therapies. On the other hand, those predicted to have longer progression-free intervals could benefit from selective de-escalation of chemotherapy to reduce toxicity.

While our results demonstrate improved prognostic accuracy through a multimodal approach, there are several limitations that must be addressed. Firstly, we use a modest sample size (n=387 total, n=78 test set) of patients treated at a single institution with carefully curated clinical features and reproducible radiomic features.^9^ Although we evaluate results on multiple feature sets, model types, and cross-validation folds, we do not achieve traditional statistical significance, likely due to the limited test size. Our results suggest that while we capture relevant prognostic signal, our study would benefit from definitive multi-institutional validation and larger, independent cohorts.

Secondly, we were unable to include more events of non-recurrence in our sample. While the observed frequency of events was consistent with the clinical trajectory of the disease and appropriate for building survival models that predict recurrence time rather than just binary risk, the methodological decision was partly influenced by a lack of follow-up data for patients being studied. We minimised retrospective selection bias through consecutive patient sampling from a defined source population with a strict inclusion criterion that formed a real-world representative cohort rather than a curated research sample.

Thirdly, the high feature-to-sample ratio, specifically for the ‘All Features’ feature set (25 features for 309 training samples), does constrain our model’s performance. However, our study is expected to have some extent, curbed the effects of this curse of dimensionality through rigorous cross-validation, our choice of tree-based models that are innately less susceptible to overfitting, and by implementing a feature selection pipeline that successfully removed a majority of redundant features. Our feature selection technique used a combination of XGBoost-based importance and univariate Cox filtering to ensure features are selected across tree-based and linear model architectures and do have survival predictive power. These features were used consistently to train all 3 model’s (XGBoost, RSF, Cox) and enabled direct performance comparison without model-specific bias. We also recognise that our model performance may benefit from adopting a late-fusion technique in future studies compared to the feature-level fusion technique that is currently used.

Furthermore, we understand that MR-derived radiomics may be affected by motion induced deterioration effects and patient intolerance of Gadolinium contrast may limit applicability across broader clinical scenarios.^42^ Further research aims to use a segmentation pipeline compatible with a diverse range of imaging techniques for wider applicability.

Lastly, our model focuses on studying first progression post radiotherapy using radiomics derived from the patient’s baseline MRI. In this study, we did not follow the changes over a longitudinal time-period from diagnosis to recurrence and/or death. Integrating this information can allow for a stronger and more clinically comprehensive model.

## Conclusion

Multimodal survival models (particularly RSF) have demonstrated prognostic value in GBM IDH-wt, and a clear improvement compared to baseline clinical models that constitute current practice. Radiomic features, specifically from the perilesional oedema and the white matter disconnectome, are novel yet can prove to be significant in imaging studies. Large-scale validation can pave the way for future translation of predictive frameworks into routine clinical decision making, enhancing personalied patient care in neuro-oncology.

## Ethical Approval

The local ethics committee at University College London approved the study. We received ethical permission for the consentless analysis of local irrevocably anonymised data collected during routine clinical care.

## Authorship Statement

*Conception and design:* MT, HH, JKR. *Data acquisition:* MT, MB, AT, HP, HH, JKR. *Analysis & interpretation:* MT, HH, JKR. *Segmentation & radiomics extraction:* JKR. *Model design, training, & implementation:* MT, JKR. *Manuscript drafting:* MT, HH, JKR. *Critical revision:* All authors. *Study supervision:* HH, JKR.

## Data Availability

Our study ethics preclude the release of internal data. Individual patient imaging and data cannot be shared to privacy regulations. The analysis code shall be openly available on publication. Model hyperparameters are provided in the supplementary materials.

## References

1. Stupp R, Mason WP, van den Bent MJ, et al. Radiotherapy plus Concomitant and Adjuvant Temozolomide for Glioblastoma. New England Journal of Medicine. 2005;352(10):987–996. Doi:10.1056/nejmoa043330

2. Louis DN, Perry A, Wesseling P, et al. The 2021 WHO Classification of Tumors of the Central Nervous System: a summary. Neuro-Oncology. 2021;23(8). Doi:10.1093/neuonc/noab106

3. Long H, Zhang P, Bi Y, Yang C, Wang J. MRI Radiomic Features of Peritumoral Edema May Predict the Recurrence Sites of Glioblastoma Multiforme. SSRN Electronic Journal. Published online January 1, 2021. Doi:10.2139/ssrn.3887443

4. Wei Y, Li C, Cui Z, et al. Structural connectome quantifies tumour invasion and predicts survival in glioblastoma patients. Brain. 2022;146(4):1714–1727. Doi:10.1093/brain/awac360

5. Alessandro Salvalaggio, Pini L, Bertoldo A, Maurizio Corbetta. Glioblastoma and brain connectivity: the need for a paradigm shift. The Lancet Neurology. 2024;23(7):740–748. Doi:10.1016/s1474-4422(24)00160-1

6. Chinot OL, Macdonald DR, Abrey LE, Zahlmann G, Kerloëguen Y, Cloughesy TF. Response Assessment Criteria for Glioblastoma: Practical Adaptation and Implementation in Clinical Trials of Antiangiogenic Therapy. Current Neurology and Neuroscience Reports. 2013;13(5). Doi:10.1007/s11910-013-0347-2

7. Ruffle JK, Samia Mohinta, Gray R, Harpreet Hyare, Parashkev Nachev. Brain tumour segmentation with incomplete imaging data. Brain communications. 2023;5(2). Doi:10.1093/braincomms/fcad118

8. Thiebaut de Schotten M, Foulon C, Nachev P. Brain disconnections link structural connectivity with function and behaviour. Nature Communications. 2020;11(1). Doi:10.1038/s41467-020-18920-9

9. van Griethuysen JJ, Fedorov A, Parmar C, et al. Computational Radiomics System to Decode the Radiographic Phenotype. Cancer research. 2017;77(21):e104–e107. Doi:10.1158/0008-5472.CAN-17-0339

10. CamDavidsonPilon. GitHub – CamDavidsonPilon/lifelines: Survival analysis in Python. GitHub. Published October 29, 2024. https://github.com/CamDavidsonPilon/lifelines

11. Chen T, Guestrin C. XGBoost: a Scalable Tree Boosting System. Proceedings of the 22nd ACM SIGKDD International Conference on Knowledge Discovery and Data Mining – KDD ‘16. 2016;1(1):785–794. Doi:10.1145/2939672.2939785

12. Pölsterl S. scikit-survival: A Library for Time-to-Event Analysis Built on Top of scikit-learn. Journal of Machine Learning Research. 2020;21(212):1–6. https://www.jmlr.org/papers/v21/20-729.html

13. Ma B, Yan G, Chai B, Hou X. XGBLC: an improved survival prediction model based on XGBoost. Bioinformatics. 2021;38(2):410–418. Doi:10.1093/bioinformatics/btab675

14. Moncada-Torres A, van Maaren MC, Hendriks MP, Siesling S, Geleijnse G. Explainable machine learning can outperform Cox regression predictions and provide insights in breast cancer survival. Scientific Reports. 2021;11(1). Doi:10.1038/s41598-021-86327-7

15. Ishwaran H, Kogalur UB, Blackstone EH, Lauer MS. Random survival forests. The Annals of Applied Statistics. 2008;2(3):841–860. Doi:10.1214/08-aoas169

16. Jain K, Jindal R. Sampling and noise filtering methods for recommender systems: A literature review. Engineering Applications of Artificial Intelligence. 2023;122:106129. Doi:10.1016/j.engappai.2023.106129

17. Wilson M. Survival Analysis II Cox Proportional Hazards Models.; 2018. https://health.ucdavis.edu/media-resources/ctsc/documents/pdfs/cph-model-presentation.pdf

18. Baldé B. Bayesian Sorcery for Hyperparameter Optimization using Optuna. Medium. Published June 9, 2023. https://medium.com/@becaye-balde/bayesian-sorcery-for-hyperparameter-optimization-using-optuna-1ee4517e89a

19. Akiba T, Sano S, Yanase T, Ohta T, Koyama M. Optuna: A Next-generation Hyperparameter Optimization Framework. arXiv:190710902 [cs, stat]. Published online July 25, 2019. https://arxiv.org/abs/1907.10902

20. Longato E, Vettoretti M, Di Camillo B. A practical perspective on the concordance index for the evaluation and selection of prognostic time-to-event models. Journal of Biomedical Informatics. 2020;108:103496. Doi:10.1016/j.jbi.2020.103496

21. Breiman L. Random Forests. Machine Learning. 2001;45(1):5–32.

22. Mangiola A, Saulnier N, De Bonis P, et al. Gene Expression Profile of Glioblastoma Peritumoral Tissue: An Ex Vivo Study. Pani G, ed. PloS ONE. 2013;8(3):e57145. Doi:10.1371/journal.pone.0057145

23. Boehm KM, Khosravi P, Vanguri R, Gao J, Shah SP. Harnessing multimodal data integration to advance precision oncology. Nature Reviews Cancer. 2022;22(2):114–126. Doi:10.1038/s41568-021-00408-3

24. Verleysen M, François D. The Curse of Dimensionality in Data Mining and Time Series Prediction. Computational Intelligence and Bioinspired Systems. Published online 2005:758-770. Doi:10.1007/11494669_93

25. Harrell F. Regression Modeling Strategies with Applications to Linear Models, Logistic and Ordinal Regression, and Survival Analysis Second Edition Springer Series in Statistics.; 2015. https://nibmehub.com/opac-service/pdf/read/Regression%20Modeling%20Strategies-%202nd%20edition-%202015.pdf

26. Xiao J, Mo M, Wang Z, et al. The Application and Comparison of Machine Learning Models for the Prediction of Breast Cancer Prognosis: Retrospective Cohort Study. JMIR Medical Informatics. 2022;10(2):e33440. Doi:10.2196/33440

27. Chen W, Zhou B, Jeon CY, et al. Machine learning versus regression for prediction of sporadic pancreatic cancer. Pancreatology. 2023;23(4):396–402. Doi:10.1016/j.pan.2023.04.009

28. Srivastava DK, George EO, Lu Z, Rai SN. Impact of unequal censoring and insufficient follow-up on comparing survival outcomes: Applications to clinical studies. Statistical methods in medical research. 2021;30(9):2057. Doi:10.1177/09622802211017592

29. Razvan Onciul, Felix-Mircea Brehar, Dumitru AV, et al. Predicting overall survival in glioblastoma patients using machine learning: an analysis of treatment efficacy and patient prognosis. Frontiers in Oncology. 2025;15. Doi:10.3389/fonc.2025.1539845

30. Yun YC, Jende JME, Garhöfer F, et al. Combined peritumoral radiomics and clinical features predict 12-month progression free survival in glioblastoma. Journal of Neuro-Oncology. Published online April 17, 2025. Doi:10.1007/s11060-025-05037-6

31. Pignotti F, Ius T, Russo R, et al. Development and validation of a MRI-radiomics-based machine learning approach in High Grade Glioma to detect early recurrence. Frontiers in Oncology. 2024;14. Doi:10.3389/fonc.2024.1449235

32. Denfeld QE, Burger D, Lee CS. Survival analysis 101: an easy start guide to analysing time-to-event data. Published online February 7, 2023. Doi:10.1093/eurjcn/zvad023

33. Turkson AJ, Ayiah-Mensah F, Nimoh V. Handling Censoring and Censored Data in Survival Analysis: A Standalone Systematic Literature Review. Tang N, ed. International Journal of Mathematics and Mathematical Sciences. 2021;2021:1–16. Doi:10.1155/2021/9307475

34. Ruiz-Ontañon P, Orgaz JL, Aldaz B, et al. Cellular Plasticity Confers Migratory and Invasive Advantages to a Population of Glioblastoma-Initiating Cells that Infiltrate Peritumoral Tissue. Stem Cells. 2013;31(6):1075–1085. Doi:10.1002/stem.1349

35. Zhang X, Zhang W, Mao XG, Zhen HN, Cao WD, Hu SJ. Targeting Role of Glioma Stem Cells for Gliobastoma Multiforme. Current Medicinal Chemistry. 2013;20(15):1974–1984. Doi:10.2174/0929867311320150004

36. Chen J, Li Y, Yu TS, et al. A restricted cell population propagates glioblastoma growth after chemotherapy. Nature. 2012;488(7412):522–526. Doi:10.1038/nature11287

37. Mangiola A, de Bonis P, Maira G, et al. Invasive tumor cells and prognosis in a selected population of patients with glioblastoma multiforme. Cancer. 2008;113(4):841–846. Doi:10.1002/cncr.23624

38. Wu CX, Lin GS, Lin ZX, et al. Peritumoral edema on magnetic resonance imaging predicts a poor clinical outcome in malignant glioma. Oncology Letters. 2015;10(5):2769–2776. Doi:10.3892/ol.2015.3639

39. Hu J, Bao H, Liu X, et al. Glioma-white matter tract interactions: A diffusion magnetic resonance imaging-based 3-tier classification and its clinical relevance. Neuro-Oncology. 2025;27(7):1888–1898. Doi:10.1093/neuonc/noaf036

40. Wen PY, Macdonald DR, Reardon DA, et al. Updated Response Assessment Criteria for High-Grade Gliomas: Response Assessment in Neuro-Oncology Working Group. Journal of Clinical Oncology. 2010;28(11):1963–1972. Doi:10.1200/jco.2009.26.3541

41. Shekarian T, Ritz MF, Hogan S, et al. Multidimensional analysis of matched primary and recurrent glioblastoma identifies contributors to tumor recurrence influencing time to relapse. Journal of neuropathology and experimental neurology. 2025;84(1):45–58. Doi:10.1093/jnen/nlae108

42. Zaitsev M, Maclaren J, Herbst M. Motion Artifacts in MRI: a Complex Problem with Many Partial Solutions. Journal of Magnetic Resonance Imaging. 2015;42(4):887–901. Doi:10.1002/jmri.24850

43. Ruffle JK, Samia Mohinta, Pombo G, et al. Brain tumour genetic network signatures of survival. Brain. 2023;146(11):4736–4754. doi:10.1093/brain/awad199

