## Supplementary Table 1 for "Predicting progression-free survival in glioblastoma: influence of the perilesional oedema and white-matter disconnectome" (from another version of this preprint: version 1, 2026-02-28)

***Supplementary Table 1:*** *Three-dimensional shape based radiomic features extracted using PyRadiomics. 17 shape features were extracted for each of the five lesion compartments: 1) full lesion (tumour + oedema), 2) full tumour (enhancing + non-enhancing tumour), 3) enhancing tumour, 4) non-enhancing tumour, 5) perilesional oedema. For each of the five compartments, all 17 features are computed for the original lesion mask and the disconnectome map, resulting in 34 radiomic features. This yields a subtotal of 170 radiomic features (17 features × 5 compartments × 2 representations (original, disconnectome)). We use two compartments for this study (full tumour and perilesional oedema).*

| Feature Name | PyRadiomics Class |
|---|---|
| **Voxel Volume** | shape |
| **Mesh Volume** | shape |
| **Surface Area** | shape |
| **Surface Volume Ratio** | shape |
| **Compactness 1** | shape |
| **Compactness 2** | shape |
| **Sphericity** | shape |
| **Spherical Disproportion** | shape |
| **Maximum 3D Diameter** | shape |
| **Maximum 2D Diameter Slice** | shape |
| **Maximum 2D Diameter Column** | shape |
| **Maximum 2D Diameter Row** | shape |
| **Major Axis Length** | shape |
| **Minor Axis Length** | shape |
| **Least Axis Length** | shape |
| **Elongation** | shape |
| **Flatness** | shape |
