## supplementary table 2 for "Predicting progression-free survival in glioblastoma: influence of the perilesional oedema and white-matter disconnectome" (from another version of this preprint: version 1, 2026-02-28)

***Supplementary Table 2:*** *Adaptive hyperparameter search spaces for all 4 feature sets (Clinical, Clinical + Tumour, Clinical + Oedema, All Features) for each of the 3 models (XGBoost, RSF, CoxPH). All parameters were sampled across 100 trials with 5-fold-cross-validation.*

| Model | Parameter | Clinical Features | Clinical + Tumour Features | Clinical + Oedema Features | All Features |
|---|---|---|---|---|---|
| **XGBoost** | Learning Rate | [0.02, 0.10] | [0.02, 0.08] | [0.008, 0.06] | [0.008, 0.08] |
| | Maximum Depth | [3, 5] | [3, 4] | [2, 3] | [3, 5] |
| | Minimum Child Weight | [4, 8] | [6, 10] | [8, 14] | [6, 14] |
| | Subsample | [0.65, 0.85] | [0.65, 0.85] | [0.55, 0.80] | [0.55, 0.85] |
| | Column sample by Tree | [0.65, 0.85] | [0.65, 0.85] | [0.55, 0.75] | [0.55, 0.85] |
| | Reg Alpha | [0.8, 5.0] | [2.0, 8.0] | [3.5, 12.0] | [1.5, 15.0] |
| | Reg Lambda | [1.0, 5.0] | [2.0, 8.0] | [3.5, 12.0] | [1.5, 15.0] |
| | Gamma | [0.5, 2.5] | [1.0, 5.0] | [2.0, 7.0] | [0.8, 8.0] |
| **RSF** | Number of Estimators | [50, 150] | [200, 600] | [150, 600] | [200, 600] |
| | Maximum Depth | [2, 3] | [3, 5] | [2, 6] | [3, 5] |
| | Minimum Samples Split | [20, 40] | [10, 30] | [10, 30] | [10, 30] |
| | Minimum Samples Leaf | [10, 30] | [8, 25] | [5, 20] | [8, 25] |
| | Maximum Features | {sqrt} | {sqrt, log2} | {sqrt, log2} | {sqrt, log2} |
| **CoxPH** | Penalizer | [0.01, 100.0] | [0.01, 5.0] | [0.01, 1.0] | [0.005, 1.0] |
| | L1 Ratio | [0.0, 1.0] | [0.0, 0.2] | [0.0, 0.7] | [0.0, 0.05] |
